# Cardiac Arrhythmias in Patients with COVID-19: A Systematic review and Meta-analysis

**DOI:** 10.1101/2020.10.09.20209379

**Authors:** Omar Hamam, Ahmed Goda, Moustafa Eldalal, Amr Ussama, Mostafa Fahmy, Karim Elyamany, Waleed Ikram, Ahmed Mahdy, Renu Bhandari, Mohammad A. Elbahnasawy, Hosam Asal, Alexander Egbe

## Abstract

**Background:** Cardiac arrhythmia cannot be overlooked in patients with coronavirus disease 2019 (COVID-19) as it carries a great influence on the outcomes. Hence, this study aimed to build concrete evidence regarding the incidence of cardiac arrhythmia in patients with COVID-19.

**Methods:** We performed a systematic search for trusted databases/search engines including PubMed, Scopus, Cochrane library and web of science. After screening, the relevant data were extracted and the incidences from the different included studies were pooled for meta-analysis.

**Results:** Nine studies were finally included in our study consisting of 1445 patients. The results of meta-analysis showed that the incidence of arrhythmia in patients with COVID-19 was 19.7% with 95% confidence interval (CI) ranging from 11.7 to 27.6%. There was also a significant heterogeneity (*I*^2^□=□94.67%).

**Conclusion:** Cardiac arrhythmias were highly frequent in patients with COVID-19 and observed in 19.7% of them. Appropriate monitoring by electrocardiogram with accurate and early identification of arrhythmias is important for better management and outcomes.

**Highlights:**

- Cardiac arrhythmia cannot be overlooked in patients with coronavirus disease 2019 (COVID-19) as it carries a great influence on the outcomes.
- This study aimed to build concrete evidence regarding the incidence of cardiac arrhythmia in patients with COVID-19.
- Cardiac arrhythmias were highly frequent in patients with COVID-19 and observed in 19.7% of them.
- Appropriate monitoring by electrocardiogram with accurate and early identification of arrhythmias is important for better management and outcomes.

## INTRODUCTION

Coronavirus disease 2019 (COVID-19) has become a global health concern affecting the lives of millions around the world^1^. It is caused by the severe acute respiratory syndrome coronavirus 2 (SARS-CoV-2). The clinical manifestations can differ from very mild to highly severe with the mean affection of the respiratory system which can lead to pneumonia as well as respiratory distress syndrome^2,3^. Moreover, the associated involvement of other systems such as the cardiovascular system cannot be overlooked as it carries a great influence on the outcomes^2-6^.

In a recent published report, cardiac involvement was showed to occur in COVID-19 even without manifestations of respiratory tract involvement^7^. Several studies asserted upon the incidence of cardiac injury in COVID-19^5,8^. Among those reported cardiac manifestations, cardiac arrhythmias were observed with variable estimates among studies and according to severity levels^2,6^.

Cardiac arrhythmias appear to be one of the highly challenging complications of COVID-19 and several speculations about different mechanisms were reported with a key role for pro-inflammatory mediators. Some of ion channels can be affected due to COVID-19 which may lead to subsequent alterations in cardiac conduction or repolarization. Thus, it can increase the risk for cardiac arrhythmogenesis to occur ^9-12^. Wang et al^2^ was the first to report an incidence of 17% of arrhythmias in COVID-19 patients. Most of those cases with arrhythmias were transferred to intensive care unit. Likewise, more and higher incidence of arrhythmias (44%) was reported in studies involving patients in the intensive care unit ^2,6^. Therefore, a better understanding of the magnitude and role of cardiac arrhythmias in COVID-19 is essential for better monitoring and management. Hence, this systematic review and meta-analysis aimed to build concrete evidence regarding the incidence of cardiac arrhythmia in COVID-19.

## METHODS

We adhered to the Preferred Reporting Items for Systematic Reviews and Meta-Analyses (PRISMA) guidelines and Cochrane’s handbook of systematic reviews to conduct this study^13,14^.

### Literature search

We combined the following keywords and conducted our search: “COVID-19”, “SARS-CoV-2”, “cardiovascular” and “Arrhythmia”. We searched PubMed, Web of Science, Scopus, and Cochrane Library for relevant articles to be included. An additional online and manual search was performed on Google Scholar and Preprint Servers to ensure adequate inclusion of all studies.

### Eligibility criteria

Results were imported into Endnote X8 (Thompson Reuter, CA, USA) for duplicates deletion. We included valid case series (>10 patients) and cohort studies including adults with COVID-19 with incidence of arrhythmia. Review articles, editorial, commentaries were excluded.

### Studies selection

The first author (O.H) divided other authors into two teams; each team independently performed title and abstract screening. Then, each team obtained the full-text of the included papers and performed full-text screening. Any disagreement between the two teams was resolved through consultation with the study senior (A.E).

### Data extraction

The two teams extracted the data; one team performed extraction of selected outcomes and the other team extracted baseline data, then, data were revised in a cross-revision manner. Extracted data include author, country, year, study design, age, sex, total number of patients, and number of patients with arrhythmia.

### Risk of bias assessment

We used the Newcastle–Ottawa scale (NOS) which is available at (https://www.ohri.ca/programs/clinical_epidemiology/oxford.asp) for assessing the risk of bias for our included studies. The possible scores of this scale range from 0 to 9. Studies with a score of seven to nine, four to six, and zero to three were classified as studies with low, moderate, and high risk of bias, respectively.

### Data synthesis and analysis

The meta-analysis of the included studies was performed using OpenMeta [Analyst] version 1.15 for conducting single-arm meta-analysis. Meta-analysis for proportions was utilized to pool the incidence of arrhythmia in the groups. Dichotomous data were calculated to obtain risk ratios along with their 95% confidence intervals (CIs). Heterogeneity among studies was assessed using the I^2^ test and P-value from the chi-squared test of heterogeneity. Values of I^2^ >50 and P<0.1 are significant markers of heterogeneity among studies according to Cochrane’s handbook^14^. Random effect models were used to avoid the effect heterogeneity. The statistical significance was set with P-value at 0.05. According to Egger and colleagues, assessment of publication bias using funnel plot method and Egger’s test is unreliable for less than 10 included studies. Therefore, in the present study, we could not assess for publication bias due to small number of included studies^15,16^.

## RESULTS

### Results of the literature search

We searched the aforementioned search engines/databases and found 955 studies after duplicate removal. We excluded 946 studies as they were not eligible for inclusion according to eligibility criteria, and a total of nine studies were finally included in our study consisting of 1445 patients^2,5,6, 7-22^. Figure (1) shows a summary of our search and table (1) shows the summary of the included studies.

**Figure 1.**
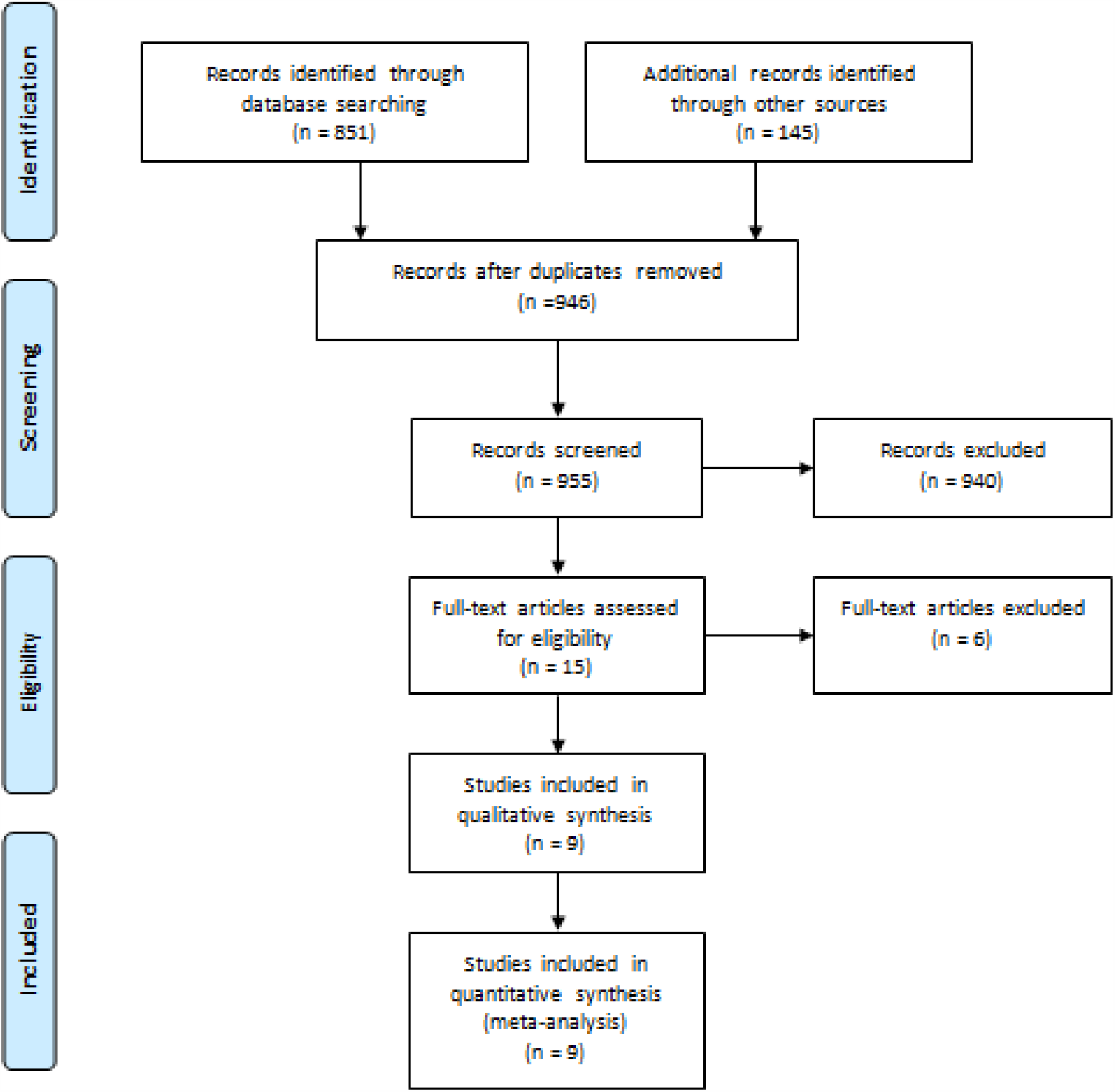
PRISMA flow diagram demonstrating the search process.

**Table 1.** Characteristics of included studies.

| Author/year of publication | Study design | Country | No. of patients | Age (Mean/median) (years) | Male % | No. of patients with cardiac arrhythmia | NOS |
| --- | --- | --- | --- | --- | --- | --- | --- |
| **Guo 2020** | Retrospective case series | China | 187 | 58.5 | 48.7 | 11 | 7 |
| **Hu 2020** | Retrospective case series | China | 323 | 61 | 51.4 | 98 | 7 |
| **Wang 2020a** | Retrospective case series | China | 138 | 56 | 54.3 | 23 | 7 |
| **Du 2020** | Retrospective case series | China | 85 | 65.8 | 72.9 | 51 | 7 |
| **Zhang 2020** | Retrospective case series | China | 221 | 55 | 48.9 | 24 | 7 |
| **Wang 2020b** | Retrospective case series | China | 339 | 71 | 49 | 35 | 7 |
| **Cao 2020** | Retrospective case series | China | 102 | 54 | 52 | 18 | 7 |
| **Lei 2020** | Retrospective case series | China | 34 | 55 | 41.2 | 8 | 8 |
| **Aggarwal 2020** | Retrospective case series | USA | 16 | 67 | 75 | 1 | 7 |

NOS, Newcastle Ottawa Scale

### Baseline characteristics

Baseline characteristics are displayed in table 1. Eight of the included studies were conducted in China while only one study reported from USA. Among the included studies, the highest mean age was 71 years while the lowest was 55 years. Most of the included studies have a male predominance reaching 75% of the total included patients.

### Risk of bias assessment

Among our nine studies evaluated for the risk of bias, all of our studies had a low risk of bias with score of seven or higher (Table 1).

### Arrhythmia in COVID-19

The results of meta-analysis including the nine studies showed that the incidence of arrhythmia in patients with COVID-19 was 19.7% with the 95% confidence interval (CI) of 11.7 to 27.6%. There was significant heterogeneity (*I*^2^ □ = □ 94.67%) which can be attributed to the variability of reported incidences among the different included studies (Figure 2).

**Figure 2.**
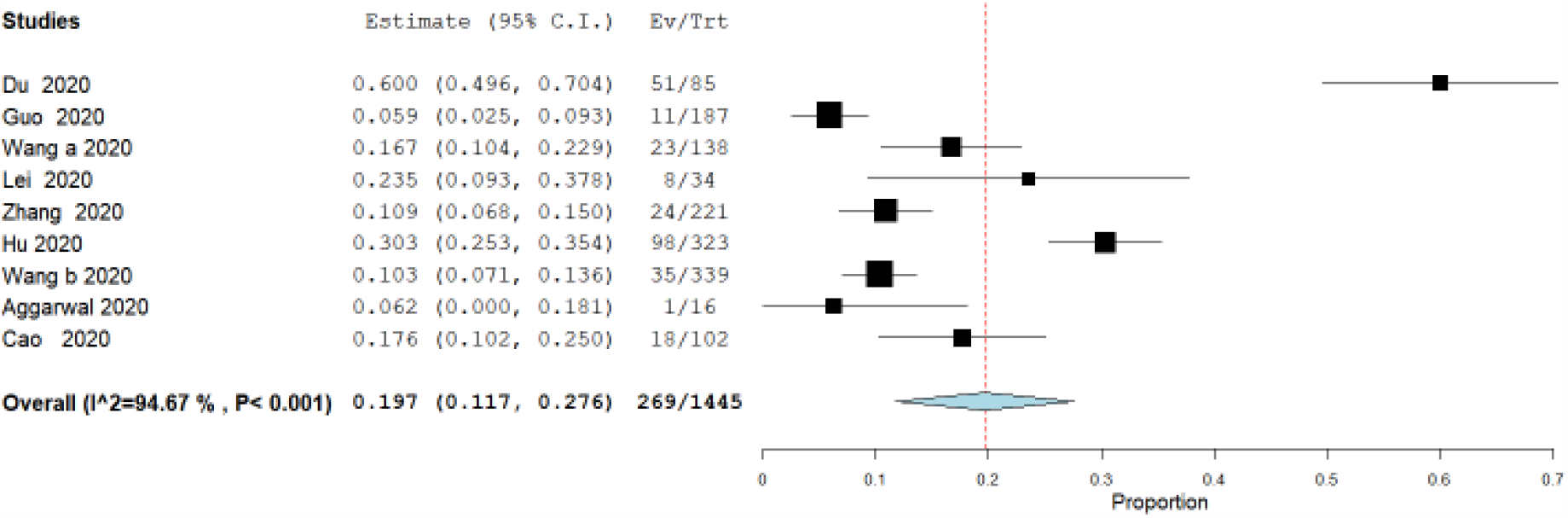
Incidence of arrhythmia in COVID-19 patients.

## DISCUSSION

Previous studies has showed that compared to different systems, the cardiovascular system is considered the second most commonly affected organ by COVID-19 after the respiratory system ^2,5-8,23^. The results of this systematic review and meta-analysis showed that cardiac arrhythmia occurred frequently in patients with COVID-19 with incidence exceeding 19% of the total cases with covid-19. This is consistent with other individual studies reported firstly from china such as Wang et al ^2^ who reported that cardiac arrhythmia occurred in 23 patients out of 138 patients included in their study. In our study, the cumulative number of patients with both arrhythmia and COVID-19 was 269 out of 1445 patients with COVID-19 which is slightly of higher percentage than Wang et al^2^. It is known from previous meta-analyses that baseline characteristics such as sex, hypertension, and diabetes can affect the outcomes of the pooled studies^23-27^ but the meta-regression option was not available to assess the exact effect of those factors given that the number of the included studies was less than 10.

There are different documented speculations to explain the association of arrhythmia with COVID-19. For instance, this may be correlated with metabolic disruption, hypoxia, pro-inflammatory process, or different stressors associated coming on the line with COVID-19 ^28^ Moreover, some of ion channels can be affected due to COVID-19 which may lead to subsequent alterations in cardiac conduction or repolarization. Thus, it can increase the risk for cardiac arrhythmia to occur^9-11^.

We noticed significant heterogeneity while performing this meta-analysis. This can be explained by the different and variable estimates among the included studies which were the highest in Du et al al. ^20^ reaching 60%. It is also worth noting that our included studies did not mention the types of arrhythmia in specific which prevent us from subgrouping the incidence for each type.

This study can be considered the most updated and comprehensive study to assess the incidence of COVID-19 in a suitable number of patients. However, this study suffered from several limitations. Although the number of patients from our pooled studies was 1445 patients, the number of studies was only nine which lead us to avoid testing for publication bias and performing meta-regression according to the reported recommendations for those tests that need at least 10 studies to be well performed. Moreover, the types of cardiac arrhythmias were not specified so we could not provide more specific subgroup analysis based upon the included studies. The studies were all observational retrospective and this type of studies has its own known limitations. Further studies that can explore cardiac arrhythmias in COVID-19 patients with more details on determinants are highly recommended.

To recapitulate, cardiac arrhythmias were highly frequent in patients with COVID-19 and observed in 19.7% of them. Appropriate monitoring by electrocardiogram with accurate and early identification of arrhythmias is important for better management and outcomes.

## Data Availability

Available upon request from the corresponding author

## Authors’ contribution

O.H. and A.G. conceived and designed the study. All authors acquired the data, performed the data extraction, and performed extensive research on the topic. A.E. reviewed and performed extensive editing of the manuscript. All authors contributed to the writing of the manuscript. O.H. performed the statistical analysis.

## Acknowledgement

None

## REFERENCES

1. World Health Organization. Coronavirus disease 2019 (COVID-19) Situation Report e 95. 2020.

2. Wang D, Hu B, Hu C, Zhu F, Liu X, Zhang J, et al. Clinical characteristics of 138 hospitalized patients with 2019 novel coronavirus-infected pneumonia in Wuhan, China. JAMA 2020;323(11):1061. https://doi.org/10.1001/jama.2020.1585.

3. Chen N, Zhou M, Dong X, Qu J, Gong F, Han Y, et al. Epidemiological and clinical characteristics of 99 cases of 2019 novel coronavirus pneumonia in Wuhan, China: a descriptive study. Lancet. 2020;395(10223):507–13.

4. Huang C, Wang Y, Li X, Ren L, Zhao J, Hu Y, et al. Clinical features of patients infected with 2019 novel coronavirus in Wuhan, China. Lancet. 2020;395(10223):497–506.

5. Guo T, Fan Y, Chen M, Wu X, Zhang L, He T, et al. Cardiovascular implications of fatal outcomes of patients with coronavirus disease 2019 (COVID-19). JAMA Cardiol 2020;e201017. https://doi.org/10.1001/jamacardio.2020.1017.

6. Cao J, Tu W-J, Cheng W, Yu L, Liu Y-K, Hu X, et al. Clinical features and shortterm outcomes of 102 patients with corona virus disease 2019 in wuhan, China. Clin Infect Dis 2020.

7. Inciardi RM, Lupi L, Zaccone G, Italia L, Raffo M, Tomasoni D, et al. Cardiac involvement in a patient with coronavirus disease 2019 (COVID-19). JAMA Cardiol 2020; https://doi.org/10.1001/jamacardio.2020.1096

8. Wang L, He W, Yu X, Liu HF, Zhou WJ, Jiang H, et al. Prognostic value of myocardial injury in patients with COVID-19. Zhong Hua Xin Xue Guan Bing Za Zhi. 2020;56:E015. https://doi.org/10.3760/cma.j.cn112148-20200313-00202

9. Chen X, Li R, Pan Z, Qian C, Yang Y, You R, et al. Human monoclonal antibodies block the binding of SARS-CoV-2 spike protein to angiotensin converting enzyme 2 receptor. Cell Mol Immunol. 2020;17(6):647–9. https://doi.org/10.1038/s41423-020-0426-7..

10. Alí A, Boutjdir M, Aromolaran AS. Cardiolipotoxicity, inflammation, and arrhythmias: role for interleukin-6 molecular mechanisms. Front Physiol. 2018;9:1866.

11. Uhler C, Shivashankar GV. Mechano-genomic regulation of coronaviruses and its interplay with ageing. Nat Rev Mol Cell Biol. 2020;21(5):247–8.

12. Zhao B, Ni C, Gao R, Wang Y, Yang L, Wei J, et al. Recapitulation of SARS-CoV-2 infection and cholangiocyte damage with human liver ductal organoids. Protein Cell. 2020;https://doi.org/10.1007/s13238-020-00718-6. [Epub ahead of print]

13. Moher D, Liberati A, Tetzlaff J, Altman DG, Altman D, Antes G, et al. Preferred reporting items for systematic reviews and meta-analyses: The PRISMA statement. PLoS Medicine. 2009.

14. Higgins JPT, Green S. Cochrane Handbook for Systematic Reviews of Interventions: Cochrane Book Series. Vol. Version 5. 2008. 1 p.

15. Terrin N, Schmid CH, Lau J, Olkin I. Adjusting for publication bias in the presence of heterogeneity. Stat Med. 2003

16. Egger M, Smith GD, Schneider M, Minder C. Bias in meta-analysis detected by a simple, graphical test. Br Med J. 1997

17. Zhang G, Hu C, Luo L, Fang F, Chen Y, Li J, et al. Clinical features and outcomes of 221 patients with COVID-19 in Wuhan, China. MedRxiv 2020. https://doi.org/10.1101/2020.03.02.20030452.

18. Hu, Ling, et al. “Risk factors associated with clinical outcomes in 323 COVID-19 hospitalized patients in Wuhan, China.” Clinical infectious diseases (2020).

19. Aggarwal S, Garcia-Telles N, Aggarwal G, Lavie C, Lippi G, Henry BM. Clinical features, laboratory characteristics, and outcomes of patients hospitalized with coronavirus disease 2019 (COVID-19): Early report from the United States. Diagnosis (Berl). 2020 May 26;7(2):91–6. PubMed PMID: 32352401. Epub 2020/05/01.

20. Du Y, Tu L, Zhu P, Mu M, Wang R, Yang P, et al. Clinical Features of 85 Fatal Cases of COVID-19 from Wuhan: A Retrospective Observational Study. Am J Respir Crit Care Med. 2020 Apr 3. PubMed PMID: 32242738.

21. Wang L, He W, Yu X, Hu D, Bao M, Liu H, et al. Coronavirus disease 2019 in elderly patients: Characteristics and prognostic factors based on 4-week follow-up. J Infect. 2020 Mar 30. PubMed PMID: 32240670. Pubmed Central PMCID: PMC7118526.

22. Lei, Shaoqing, et al. “Clinical characteristics and outcomes of patients undergoing surgeries during the incubation period of COVID-19 infection.” EClinicalMedicine (2020): 100331.

23. Momtazmanesh, Sara, et al. “Cardiovascular disease in COVID-19: a systematic review and meta-analysis of 10,898 patients and proposal of a triage risk stratification tool.” The Egyptian Heart Journal 72.1 (2020): 1–17.

24. Huang I, Lim MA, Pranata R. Diabetes mellitus is associated with increased mortality and severity of disease in COVID-19 pneumonia e a systematic review, meta-analysis, and meta-regression. Diabetes Metab Syndr Clin Res Rev 2020;14:395e403. https://doi.org/10.1016/j.dsx.2020.04.018.

25. Pranata R, Lim MA, Huang I, Raharjo SB, Lukito AA. Hypertension is associated with increased mortality and severity of disease in COVID-19 pneumonia: a systematic review, meta-analysis and meta-regression. J Renin-AngiotensinAldosterone Syst JRAAS 2020;21:147032032092689. https://doi.org/10.1177/1470320320926899.

26. Huang I, Pranata R. Lymphopenia in severe coronavirus disease-2019 (COVID19): systematic review and meta-analysis. J Intensive Care 2020;8:36. https://doi.org/10.1186/s40560-020-00453-4.

27. Driggin E, Madhavan MV, Bikdeli B, Chuich T, Laracy J, Bondi-Zoccai G, et al. Cardiovascular considerations for patients, health care workers, and health systems during the coronavirus disease 2019 (COVID-19) pandemic. J Am Coll Cardiol 2020. https://doi.org/10.1016/j.jacc.2020.03.03

